# Maternal prenatal stress is associated with altered placental microstructure in low-risk pregnancies and pregnancies with Congenital Heart Disease

**DOI:** 10.64898/2026.03.02.26347408

**Authors:** Alexandra F. Bonthrone, Daniel Cromb, Sarah Ahmad Javed, Jordina Aviles Verdera, Kuberan Pushparajah, Mary Rutherford, Jana Hutter, Serena J Counsell

**Author notes:** Corresponding Author: Dr Daniel Cromb, Centre for the Developing Brain, Research Department of Early Life Imaging, King’s College London, St Thomas’ Hospital, 1st floor South Wing, SE1 7EH. Joint primary authors (Alexandra F. Bonthrone, Daniel Cromb).

## Abstract

**Objectives:** To assess if maternal stress is higher in pregnancies with congenital heart disease (CHD) compared to low-risk pregnancies and if maternal stress is associated with placental microstructure and function. To explore if CHD alters the relationship between maternal stress and placental measures.

**Methods:** In this prospective observational study, 27 participants carrying a fetus with CHD and 42 participants with typical low-risk pregnancies underwent 1-2 combined diffusion□T2* relaxation placental MRIs from 20 weeks gestation (GA) and completed the Edinburgh Postnatal Depression Scale and State Trait Anxiety Inventory [43 male fetuses, median (IQR) GA at assessment 30.86 weeks (27.43-34.00), interval between assessments 6.00 weeks (4.86-7.14)]. 98 complete placental MRI and maternal stress datasets were available. Generalized Estimating Equations were used for analyses.

**Results:** Higher trait anxiety was associated with higher placental apparent diffusion coefficient (p=0.023) adjusting for CHD, sex, GA at assessment, GA at assessment^2^, state anxiety, depressive symptoms and previous mental health treatment. Maternal state anxiety (p=0.005) and depressive symptoms (p=0.046) were higher in pregnancies with CHD adjusting for GA at assessment and previous mental health treatment. CHD did not alter these relationships (p>0.119).

**Conclusions:** Maternal proneness to anxiety, measured with the trait anxiety inventory, is associated with increased diffusivity in the placenta, which may reflect altered microstructural maturation. Mothers with fetal CHD show more depressive symptoms and feelings of anxiety and may benefit from screening for elevated maternal stress. The findings contribute to a growing body of research regarding the influence of prenatal stress on placental development.

**Highlights:**

- Maternal stress and placental MRI data acquired in pregnancies with and without CHD
- Maternal trait anxiety is associated with increased placental diffusivity
- Maternal state anxiety and depressive symptoms are higher in fetal CHD
- State anxiety and depressive symptoms not associated with placental MRI measures
- CHD did not moderate relationships between placental MRI measures and stress

## Introduction

Maternal prenatal stress impacts over 20% of pregnancies, and is associated with poorer pregnancy and neonatal outcomes, along with long-term neurodevelopmental and physical effects on offspring^1,2^. The mechanism through which maternal stress influences pregnancy and offspring outcomes is not clear.

The placenta supports fetal oxygen and nutrient delivery, waste removal, endocrine, metabolic and immune functions. Placental development is essential to healthy fetal development and disorders of placental structure and function are associated with adverse pregnancy, neonatal and longer-term outcomes^3–6^. Prenatal maternal stress is associated with altered placental gene expression^7^, enzymatic activity^8,9^, DNA methylation^10^ and increased placental weight and thickness at birth^11,12^. However, most studies assess the placenta after delivery, when key structural and functional properties may have changed ^13^.

Placental MRI facilitates safe, non-invasive assessment of the placenta throughout gestation. Saeed and colleagues reported that during the Covid-19 pandemic, placental thickness was increased and textural features were altered in pregnancies without known virus exposure compared to pre-pandemic controls, with higher maternal stress mediating this relationship^14^. T2*-relaxometry is an indirect proxy for placental oxygenation^15,16^, while diffusion MRI provides information about tissue microstructure^17^. Combined diffusion-relaxation MRI techniques enabling the acquisition of multi-modal data in a single MR scan have been applied successfully in the placenta^18^ and have identified altered placental development prior to preterm birth and in pre-eclampsia^19,20^.

Maternal stress is higher in pregnancies with fetal congenital heart disease (CHD)^21,22^ and placental development is also altered in this population. Several studies report reduced placental weight or volume in CHD^23–26^, and recent MRI work has shown altered placental function and microstructure^27^. Gross morphological changes, such as abnormal umbilical cord insertion, are reported^28^, and placental malperfusion lesions and vascular changes identified using histopathology are a recognised feature in CHD^25,29,30^. These placental differences may contribute to impaired fetal development in CHD^31,32^. Early pre-eclampsia is associated with a six-fold increased risk of CHD in future pregnancies^33^, suggesting shared or interacting pathways. Better understanding of altered placentation in CHD and identifying the underlying mechanisms is a key focus for research in this field^34–36^, which may help explain variation in fetal development and guide monitoring during high-risk pregnancies.

The objectives of this study were to assess whether (i) maternal stress is higher in pregnancies complicated by CHD compared to low-risk pregnancies; (ii) maternal stress is associated with in-vivo placental microstructure and function. We also conducted an exploratory analysis to assess if CHD moderates the relationship between maternal stress and placental measures.

## Methods

### Ethical approval

This study was approved by the NHS Wales Research Ethics Committee [NHS REC 21/WA/0075]

### Recruitment

83 pregnant participants were recruited to the Congenital Heart Disease Imaging Programme (CHIP), a study investigating fetal brain development and placental function in a mixed cohort of typical fetuses and fetuses with CHD. Control participants experiencing a low-risk pregnancy, with the absence of pregnancy-induced hypertension (PIH), preeclampsia (PE), fetal growth restriction (FGR), or gestational diabetes (GD) at the time of enrolment, were recruited after their antenatal booking or screening appointments. Participants with a fetus with a congenital defect of the heart or great vessels suspected or confirmed on fetal echocardiography, were recruited from the fetal cardiology clinic. Participants from both the CHD and control groups who went on to receive a diagnosis of PIH, PE, FGR, GD, or where the fetus went on to deliver before 37.00 weeks gestation were excluded. Control participants where the fetus had confirmed genetic or brain abnormalities (e.g. ventriculomegaly) were also excluded. All participants were invited to have up to two fetal MRI scans from a gestational age (GA) of 20 weeks.

### MRI Acquisition and Image Processing

130 placental MRI scans were acquired on a 3T Phillips Achieva scanner with a 32 channel surface coil using previously described protocols^18,37^. Briefly, combined T2*-diffusion placental data was acquired at 3mm^3^ isotropic resolution, at multiple echo-times and diffusion-weightings, enabling simultaneous assessment of placental function and microstructure. Full details of the MR imaging parameters are in *Supplementary Table 1*. Masks of the placental parenchyma were automatically generated using an in-house nn-Unet based network, trained and validated on 4,086 3T MRI diffusion datasets^38^ (*Supplemental Figure 1*). Mean T2* relaxation time was calculated within the mask as a measure placental function (T2*). Mean apparent diffusion coefficient (ADC) was calculated using MRtrix3.

### Maternal stress measures

Maternal psychological well-being was assessed at time of MRI using two validated, standardised self-report questionnaires: the Edinburgh Postnatal Depression Scale (EPDS)^39^, and the State-Trait Anxiety Inventory (STAI)^40,41^. The EPDS is a 10-item screening tool for perinatal depression (total score range 0–30) which is validated for use during pregnancy^42^. A score of 11 or more on the EPDS shows the most sensitivity and specificity when screening for pre and postnatal depressive symptoms^43^. The STAI is a 40-item assessment which provides a measure of state anxiety, i.e. the current level of anxiety (score range 20-80), and trait anxiety, i.e. how prone an individual is to anxiety (score range 20-80). A score greater than 40 is commonly used to identify high anxiety in pregnancy^44^. Participants also stated whether they had previously received treatment for a mental health problem (yes/no). Datasets were excluded if maternal stress data was incomplete. All participants in the CHD group were seen and counselled by fetal cardiologist and fetal nurse counsellor as part of standard clinical care.

### Statistical Analysis

All statistical analyses were conducted in R v.4.4.3. Continuous variables were summarised with medians and interquartile ranges, and categorical variables using frequencies and percentages.

Generalized Estimating Equation (GEE) analyses implemented in the geepack package^45–47^ were used to examine the associations between maternal anxiety and depression, CHD diagnosis and placental development. GEE is a semiparametric regression approach that accounts for correlational structures within data, facilitating the inclusion of longitudinal data. To assess if maternal stress measures were higher in infants with CHD, GA at scan (when questionnaires were completed) was included as a covariate:

- EPDS maternal depression score/STAI state anxiety score/STAI trait anxiety score ∼ GA at scan + previous mental health treatment + CHD

Maternal stress measures were not collinear (Pearson’s r 0.698-0.753) and therefore included in one model to assess the relationship between maternal stress and placental microstructure and function. CHD status, male sex, previous mental health treatment, GA at scan and the 2nd order polynomial of GA at scan were included as covariates:

- Placental ADC/Placental T2* ∼ EPDS maternal depression score + STAI state anxiety score + STAI trait anxiety score + male sex + GA at scan + GA at scan^2^ + previous mental health treatment + CHD

To assess the moderating effect of CHD on the relationship between maternal stress and placental measures, interaction terms were introduced into the equations above:

- Placental ADC/Placental T2* ∼ EPDS maternal depression score*CHD + STAI state anxiety score + STAI trait anxiety score + male sex + GA at scan + GA at scan^2^ + previous mental health treatment
- Placental ADC/Placental T2* ∼ STAI state anxiety score*CHD + EPDS maternal depression score + STAI trait anxiety score + male sex + GA at scan + GA at scan^2^ + previous mental health treatment
- Placental ADC/Placental T2* ∼ STAI trait anxiety score*CHD + EPDS maternal depression score + STAI state anxiety score + male sex + GA at scan + GA at scan^2^ + previous mental health treatment

Based on quasi-likelihood under the independence model criterion, GEEs were fit with an exchangeable correlation structure. Holm family-wise error rate correction was used to adjust for multiple comparisons (reported as p_FWE_).

### Data availability

The processed data for this study is available upon reasonable request to the corresponding author.

## Results

### Participant Demographics

98 (36 CHD) T2*-ADC multi-echo gradient-echo MRI and maternal mental health datasets from 69 participants (27 CHD) were included in this analysis (*Table 1*, *See Supplemental Figure 2 for participant exclusions*).

**Table 1.**
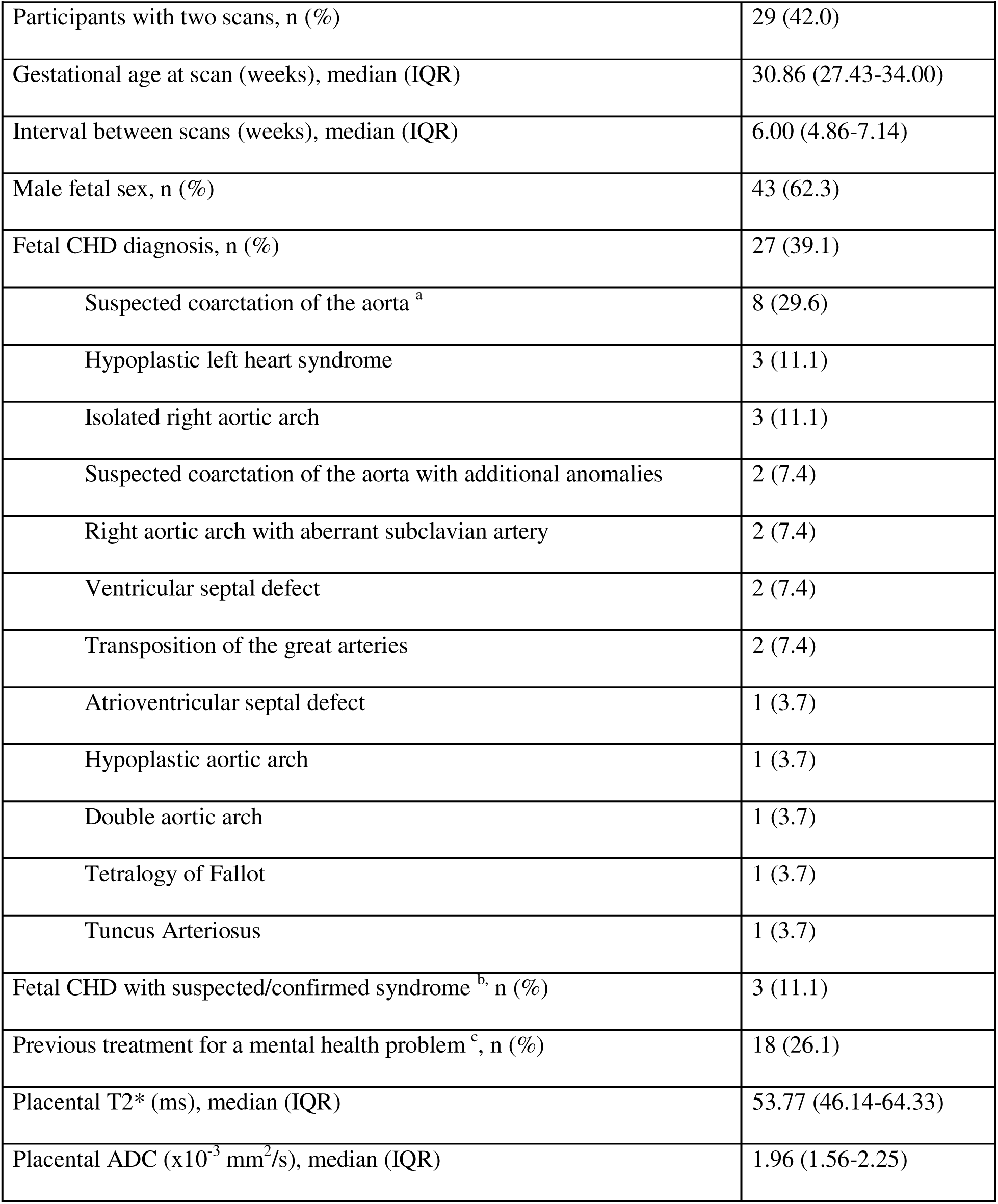
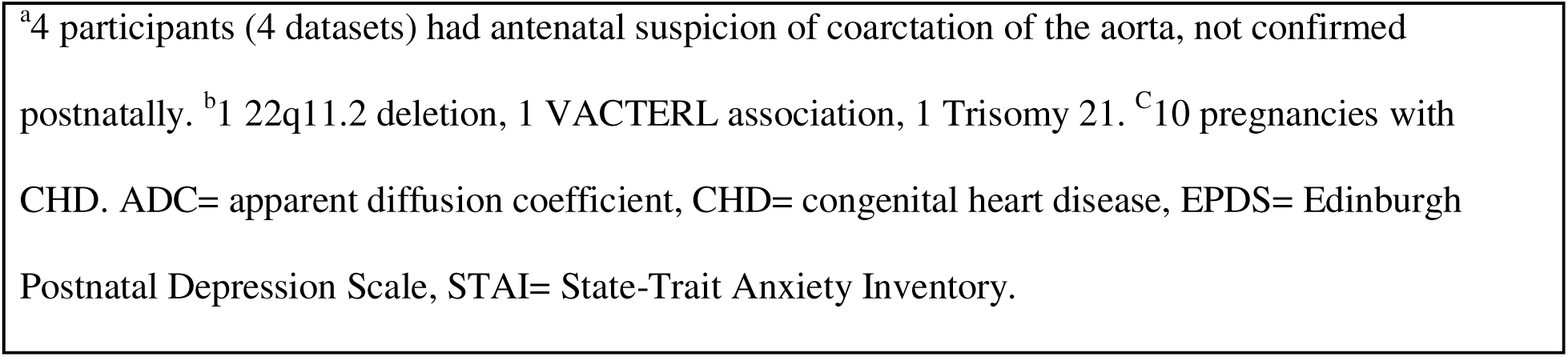
Summary of demographic, maternal stress and placental MRI data (n=69).

|  |  |
| --- | --- |
| Participants with two scans, n (%) | 29 (42.0) |
| Gestational age at scan (weeks), median (IQR) | 30.86 (27.43-34.00) |
| Interval between scans (weeks), median (IQR) | 6.00 (4.86-7.14) |
| Male fetal sex, n (%) | 43 (62.3) |
| Fetal CHD diagnosis, n (%) | 27 (39.1) |
| Suspected coarctation of the aorta <sup>a</sup> | 8 (29.6) |
| Hypoplastic left heart syndrome | 3 (11.1) |
| Isolated right aortic arch | 3 (11.1) |
| Suspected coarctation of the aorta with additional anomalies | 2 (7.4) |
| Right aortic arch with aberrant subclavian artery | 2 (7.4) |
| Ventricular septal defect | 2 (7.4) |
| Transposition of the great arteries | 2 (7.4) |
| Atrioventricular septal defect | 1 (3.7) |
| Hypoplastic aortic arch | 1 (3.7) |
| Double aortic arch | 1 (3.7) |
| Tetralogy of Fallot | 1 (3.7) |
| Tuncus Arteriosus | 1 (3.7) |
| Fetal CHD with suspected/confirmed syndrome <sup>b</sup>, n (%) | 3 (11.1) |
| Previous treatment for a mental health problem <sup>c</sup>, n (%) | 18 (26.1) |
| Placental T2* (ms), median (IQR) | 53.77 (46.14-64.33) |
| Placental ADC ($\times 10^{-3}$ mm<sup>2</sup>/s), median (IQR) | 1.96 (1.56-2.25) |

<sup>a</sup>4 participants (4 datasets) had antenatal suspicion of coarctation of the aorta, not confirmed postnatally. <sup>b</sup>1 22q11.2 deletion, 1 VACTERL association, 1 Trisomy 21. <sup>c</sup>10 pregnancies with CHD. ADC= apparent diffusion coefficient, CHD= congenital heart disease, EPDS= Edinburgh Postnatal Depression Scale, STAI= State-Trait Anxiety Inventory.

### Maternal Stress in CHD and controls

Maternal depressive symptoms and state anxiety were higher in pregnancies complicated by CHD compared to controls (*Table 2*). Two (7.4%) participants carrying a fetus with CHD [one suspected coarctation of the aorta (CoA), one hypoplastic left heart syndrome (HLHS) at two timepoints] and two (4.8%) control participants scored 11 or more on the EPDS. 10 (37%) CHD participants [4 CoA, 2 CoA with additional anomalies, one isolated right aortic arch (RAA), one RAA with aberrant subclavian artery, one HLHS, one atrioventricular septal defect (AVSD)] and six (14.3%) control participants (one at two time points) scored over 40 on the state anxiety inventory. In total, 10 (37.0%) participants carrying a fetus with CHD and seven (16.7%) controls reported elevated depressive or state anxiety symptoms. Of 17 participants with elevated symptoms, five (three CHD, two control) reported previous treatment for a mental health problem. Five (18.5%) participants with a pregnancy complicated by CHD (3 CoA, one CoA with additional anomalies, one AVSD) and five (11.9%) control participants scored over 40 on the trait anxiety inventory. Of 10 participants with elevated trait anxiety, two (both CHD) reported previously receiving treatment for a mental health problem.

**Table 2.** Maternal stress in pregnancies complicated by CHD and controls.

|  |  |  |  |  |  |
| --- | --- | --- | --- | --- | --- |
| Maternal Stress Measure | CHD<br>Median (IQR) | control<br>Median (IQR) | $\beta$ [95 % CI] | Wald<br>statistic | P-value<br>($p_{FWE}$) |
| EPDS maternal depression | 6.00<br>(2.00-9.00) | 4.00<br>(1.00-6.00) | 0.574<br>[0.079- 1.07] | 5.16 | 0.023<br>(0.046)* |
| STAI state anxiety | 35.50<br>(29.00-41.00) | 28.50<br>(23.00-35.75) | 0.774<br>[1.25- 0.295] | 10.0 | 0.002<br>(0.005)* |
| STAI trait anxiety | 33.50<br>(29.50-38.00) | 30.00<br>(24.25-37.75) | 0.432<br>[-0.064- 0.928] | 2.91 | 0.088<br>(0.088) |
| Analyses adjusted for gestational age at scan (when maternal stress was measured) and previous mental health treatment. EPDS= Edinburgh Postnatal Depression Scale, STAI= State-Trait Anxiety Inventory. *$p_{FWE}<0.05$ | | | | | |

### Maternal Stress and Placental Development

Higher maternal trait anxiety scores were associated with increased placental diffusivity (β [95 % CI] 0.323 [0.103-0.542], p_FWE_=0.023, *Figure 1*). No other significant associations were found between any of the maternal stress measures and placental T2* or ADC (*Table 3*).

**Figure 1.**
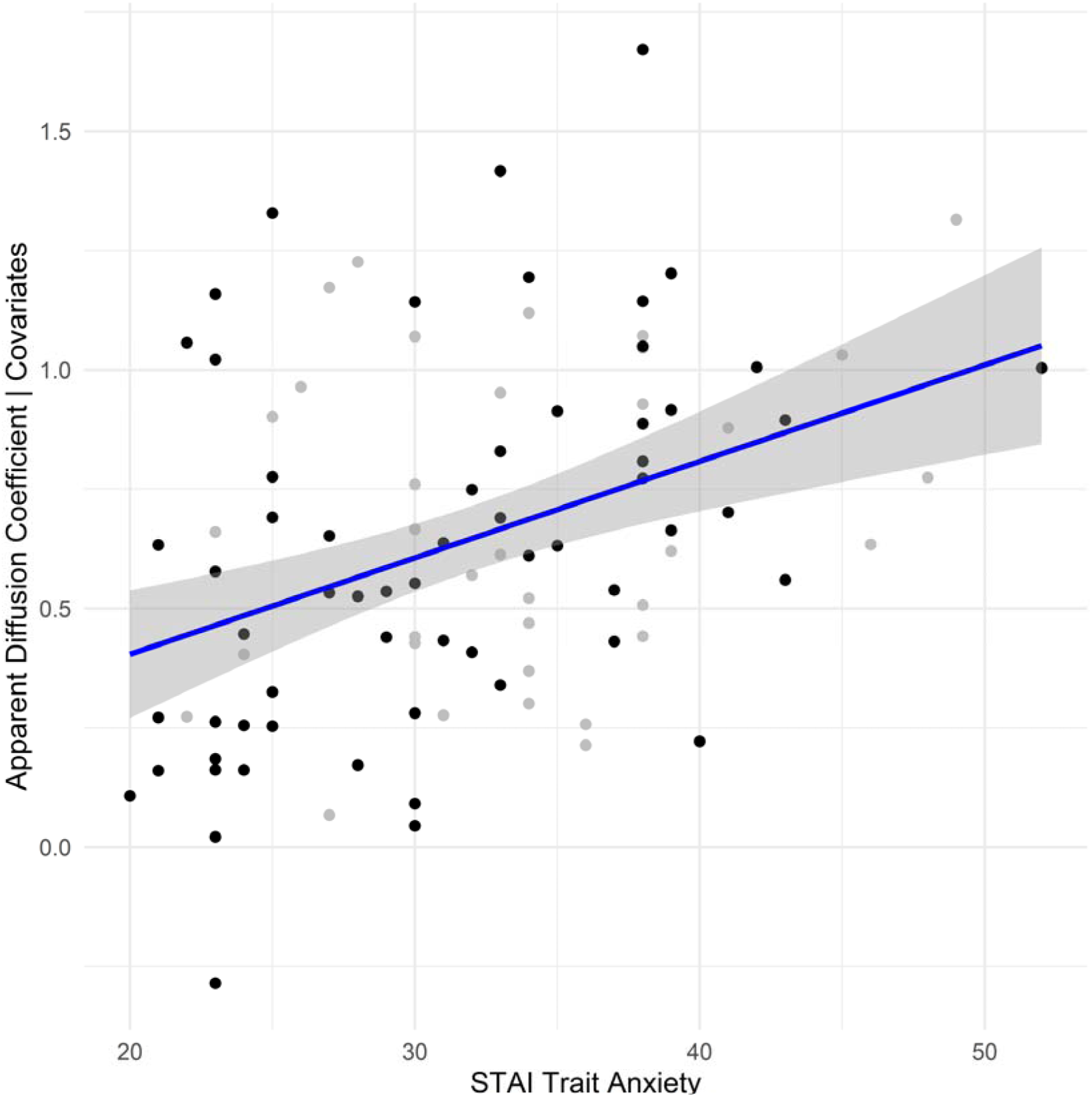
Relationship between STAI trait anxiety and placental ADC adjusted GA at scan, GA at scan^2^, male fetal sex, previous mental health treatment, CHD, STAI state anxiety and EPDS scores, grey=CHD, black=control.

**Table 3.** Associations between maternal stress measures and placental microstructure and function.

|  |  |  |  |
| --- | --- | --- | --- |
| Maternal Stress Measure | $\beta$ [95 % CI] | Wald statistic | P-value ($p_{FWE}$) |
| Placental ADC |  |  |  |
| EPDS maternal depression | -0.034 [-0.237- 0.169] | 0.110 | 0.743 (1.00) |
| STAI state anxiety | -0.211 [-0.474- 0.052] | 2.47 | 0.116 (0.581) |
| STAI trait anxiety | 0.323 [0.103- 0.542] | 8.33 | 0.004 (0.023)* |
| Placental T2* |  |  |  |
| EPDS maternal depression | 0.035 [-0.121- 0.191] | 0.200 | 0.658 (1.00) |
| STAI state anxiety | -0.037 [-0.209- 0.134] | 0.180 | 0.670 (1.00) |
| STAI trait anxiety | -0.115 [-0.313- 0.082] | 1.29 | 0.255 (1.00) |
| Analyses adjusted for presence of CHD, male sex, previous mental health treatment, GA at scan, GA at scan<sup>2</sup>. ADC= Apparent diffusion coefficient, EPDS= Edinburgh Postnatal Depression Scale, STAI= State-Trait Anxiety Inventory |  |  |  |

### Moderating effect of CHD on effect of maternal stress on placental measures

After correction for multiple comparisons, there was no moderating effect of CHD on the relationship between maternal stress measures and placental measures (p_FWE_>0.119, *Supplementary Table 2*).

## Discussion

### Main Findings

This study aimed to investigate whether maternal stress during pregnancy is associated with placental microstructure and function in a sample of low-risk pregnancies and pregnancies affected by fetal CHD. Higher maternal trait anxiety was associated with increased placental ADC, suggesting a link between proneness to anxiety and altered placental microstructural development. Maternal state anxiety and depressive symptoms were higher in mothers with CHD affected pregnancies compared to low-risk pregnancies. However, a diagnosis of fetal CHD did not moderate the relationship between maternal stress measures and placental metrics.

Higher maternal trait anxiety was associated with increased placental diffusivity across both low-risk pregnancies and pregnancies affected by fetal CHD. Previously, Saeed and colleagues reported that prenatal psychological distress mediated the association between pregnancy during the Covid-19 pandemic, but without known virus exposure, and increased placental thickness and altered textural features^14^. These findings align with previous research linking maternal stress to altered placental physiology^8–10^ and increased placental thickness^11,12^. We add to this literature by identifying associations between maternal anxiety proneness and placental microstructure. Increased maternal trait anxiety is also associated with altered placental 11β-HSD2 gene expression, which regulates fetal exposure to maternal cortisol^7,50,51^. Increased fetal cortisol exposure is associated with long-term cascading effects on neurodevelopment and health^34,37,52^. A growing body of literature that suggests placental development may be a key mediator of the relationship between maternal prenatal stress and offspring outcomes.

The biological underpinnings of the relationship between placental ADC and trait anxiety are unclear. Several cross-sectional studies report reductions in placental ADC with increasing GA^17,37,53,54^ although there is variability in ADC change over gestation in typical low-risk pregnancies^37^. Maternal stress has been linked with less heterogeneity in placental villous barrier thickness assessed postnatally^55^. The placental villous barrier becomes more heterogeneous and terminal villi vessel diameters decrease as gestation progresses. It is possible that higher placental ADC reflects larger terminal villi vessels. However, further research incorporating mental health measures, placental MRI, and histopathology are required to test this hypothesis.

We report that maternal depressive symptoms and anxiety levels were higher in pregnancies complicated by fetal CHD compared to controls. 37% of participants carrying a fetus with CHD reported elevated anxiety or depressive symptoms. This is in keeping with previous research reporting that maternal state anxiety and depressive symptoms are higher in pregnancies with fetal CHD^21,22^ and similar to pregnancies affected by preterm premature rupture of membranes (PPROM)^56^. STAI state anxiety scores in pregnancies with fetal CHD are similar to those seen in parents of infants born preterm^57^. However, only 30% of those with elevated symptoms reported previous treatment for a mental health problem, suggesting fetal CHD may be an independent risk factor for elevated maternal stress. All pregnancies complicated with fetal CHD may therefore benefit from screening for elevated maternal stress. High prenatal maternal stress can have consequences on pregnancy, neonatal and long-term offspring outcomes. There is evidence of unique associations between increased maternal prenatal stress and altered hippocampal and cerebellar volumes in fetuses with CHD compared to typical fetuses^58^. Taken together, evidence suggests that interventions are needed to minimise maternal stress in pregnancies with fetal CHD, which is in alignment with a recent statement regarding maternal perinatal mental health from the American Heart Association^59^.

### Limitations

This study has some limitations that are important to note. The CHD sample was not large (36 scans from 27 pregnancies) and therefore our investigation of the moderating effect of CHD on relationships between placental MRI measures and stress should be considered exploratory. Further research with larger samples is needed to fully characterise maternal stress and its impact on placental development in CHD. We assessed the influence of maternal stress on placental development across the 2nd half of gestation. However, stress exposure at different periods of gestation may have distinct effects on placental development, potentially reflecting sensitive periods for intervention. It was beyond the scope of this study to determine when placental changes associated with stress occur during gestation. We also did not include any health or neurodevelopmental outcome measures in this study. Larger longitudinal studies are required to fully characterise the relationship between prenatal stress, placental structure and function, and offspring mental and physical health.

## Conclusion

Our study was the first to assess the relationship between prenatal stress, fetal CHD diagnosis, and in-vivo placental microstructure and function measured with advanced MRI. Depressive symptoms and feelings of anxiety are higher in pregnancies complicated by fetal CHD, suggesting mothers carrying fetuses with CHD should be screened for elevated maternal stress. We provide novel evidence that maternal proneness to anxiety, measured with the trait anxiety inventory, is associated with increased diffusivity in the placenta, which may reflect altered microstructural maturation. Placental microstructural development may be a key mediator of the relationship between elevated maternal stress and worse obstetric, neonatal and long-term offspring outcomes. A fetal CHD diagnosis does not alter the relationship between anxiety proneness and placental microstructural development. The findings contribute to a growing body of research regarding the influence of prenatal stress on placental development. Further research is required to determine if placental microstructure mediates the relationship between prenatal stress and maternal obstetric and offspring health and wellbeing.

## Data Availability

The processed data for this study is available upon reasonable request to the corresponding author.

## Acknowledgements and funding

We are extremely grateful to all participants who took part in this study. We are also grateful to our research radiographers, the staff at the research department of early life imaging and, the Fetal Cardiology department at the Evelina London Children’s Hospital, in particular Carole Morland. This work was funded by the Medical Research Council UK (MR/V002465/1). This work was supported by core funding from the Wellcome/EPSRC Centre for Medical Engineering (WT203148/Z/16/Z), the NIH Human Placenta Project [1U01HD087202-01], EPSRC grant EP/V034537/1, and by the National Institute for Health Research (NIHR) Biomedical Research Centre based at Guy’s and St Thomas’ NHS Foundation Trust and King’s College London and the National Institute for Health Research (NIHR) Biomedical Research Centre at University College London Hospitals NHS Foundation Trust and University College London. JH was also supported by a Wellcome Trust Sir Henry Wellcome Fellowship [201374/Z/16/Z], a UKRI FLF [MR/T018119/1] and a DFG Heisenberg professorship [502024488]. The views expressed are those of the authors and not necessarily those of the NHS, the NIHR or the Department of Health. The funders had no role in conducting research or manuscript preparation.

## Contribution to Authorship

AFB: methodology, formal analysis, writing – original draft, writing – review and editing, visualisation; DC: methodology, formal analysis, data curation, writing - original draft, writing - review and editing; SAJ: data curation, writing - review and editing; JAV: methodology, software, writing - review and editing; KP: resources, writing - review and editing; MR: resources, methodology, writing - review and editing; JH methodology, software, writing – review and editing; SJC conceptualisation; methodology; investigation; resources; data curation; writing – review and editing; supervision; funding acquisition.

## References

1. Van den Bergh BRH, van den Heuvel MI, Lahti M, Braeken M, de Rooij SR, Entringer S, et al. Prenatal developmental origins of behavior and mental health: The influence of maternal stress in pregnancy. Neurosci Biobehav Rev. 2020;117:26–64.

2. Yang F, Janszky I, Roos N, Li J, László KD. Prenatal Exposure to Severe Stress and Risks of Ischemic Heart Disease and Stroke in Offspring. JAMA Netw Open. 2023 Dec 1;6(12):e2349463.

3. Sun C, Groom KM, Oyston C, Chamley LW, Clark AR, James JL. The placenta in fetal growth restriction: What is going wrong? Placenta. 2020 July;96:10–8.

4. Koulouraki S, Paschos V, Pervanidou P, Christopoulos P, Gerede A, Eleftheriades M. Short- and Long-Term Outcomes of Preeclampsia in Offspring: Review of the Literature. Child Basel Switz. 2023 May 1;10(5):826.

5. Yang C, Baker PN, Granger JP, Davidge ST, Tong C. Long-Term Impacts of Preeclampsia on the Cardiovascular System of Mother and Offspring. Hypertens Dallas Tex 1979. 2023 Sept;80(9):1821–33.

6. Ibrahim N, Weissgold SA, Brink L, Mahgoub I, Carter B, Sethna V, et al. Examining the association between placental malperfusion assessed by histopathological examination and child and adolescent neurodevelopment: a systematic review. J Child Psychol Psychiatry. 2025 Oct;66(10):1606–20.

7. O’Donnell KJ, Bugge Jensen A, Freeman L, Khalife N, O’Connor TG, Glover V. Maternal prenatal anxiety and downregulation of placental 11β-HSD2. Psychoneuroendocrinology. 2012 June;37(6):818–26.

8. St-Pierre J, Laurent L, King S, Vaillancourt C. Effects of prenatal maternal stress on serotonin and fetal development. Placenta. 2016 Dec;48 Suppl 1:S66–71.

9. Ruffaner-Hanson C, Noor S, Sun MS, Solomon E, Marquez LE, Rodriguez DE, et al. The maternal-placental-fetal interface: Adaptations of the HPA axis and immune mediators following maternal stress and prenatal alcohol exposure. Exp Neurol. 2022 Sept;355:114121.

10. Monk C, Lugo-Candelas C, Trumpff C. Prenatal Developmental Origins of Future Psychopathology: Mechanisms and Pathways. Annu Rev Clin Psychol. 2019 May 7;15:317–44.

11. Tegethoff M, Greene N, Olsen J, Meyer AH, Meinlschmidt G. Maternal Psychosocial Stress during Pregnancy and Placenta Weight: Evidence from a National Cohort Study. PLOS ONE. 2010 Dec 31;5(12):e14478.

12. Shah RG, Salafia CM, Girardi T, Rukat C, Brunner J, Barrett ES, et al. Maternal affective symptoms and sleep quality have sex-specific associations with placental topography. J Affect Disord. 2024 Sept 1;360:62–70.

13. Nelson DM, Myatt L. The Human Placenta in Health and Disease. Obstet Gynecol Clin North Am. 2020 Mar;47(1):xv–xviii.

14. Saeed H, Lu YC, Andescavage N, Kapse K, Andersen NR, Lopez C, et al. Influence of maternal psychological distress during COVID-19 pandemic on placental morphometry and texture. Sci Rep. 2023 May 10;13(1):7374.

15. Hutter J, Jackson L, Ho A, Pietsch M, Story L, Chappell LC, et al. T2* relaxometry to characterize normal placental development over gestation in-vivo at 3T. Wellcome Open Res. 2019 Nov 5;4:166.

16. Sørensen A, Hutter J, Seed M, Grant PE, Gowland P. T2*-weighted placental MRI: basic research tool or emerging clinical test for placental dysfunction? Ultrasound Obstet Gynecol. 2020;55(3):293–302.

17. Slator PJ, Hutter J, McCabe L, Gomes ADS, Price AN, Panagiotaki E, et al. Placenta microstructure and microcirculation imaging with diffusion MRI. Magn Reson Med. 2018;80(2):756–66.

18. Hutter J, Slator PJ, Christiaens D, Teixeira RPAG, Roberts T, Jackson L, et al. Integrated and efficient diffusion-relaxometry using ZEBRA. Sci Rep. 2018 Oct 11;8(1):15138.

19. Slator PJ, Hutter J, Palombo M, Jackson LH, Ho A, Panagiotaki E, et al. Combined diffusion-relaxometry MRI to identify dysfunction in the human placenta. Magn Reson Med. 2019 July;82(1):95–106.

20. Hall M, Suff N, Slator P, Rutherford M, Shennan A, Hutter J, et al. Placental multimodal MRI prior to spontaneous preterm birth <32□weeks’ gestation: An observational study. BJOG Int J Obstet Gynaecol. 2024 Dec;131(13):1782–92.

21. Rychik J, Donaghue DD, Levy S, Fajardo C, Combs J, Zhang X, et al. Maternal psychological stress after prenatal diagnosis of congenital heart disease. J Pediatr. 2013 Feb;162(2):302–307.e1.

22. Wu Y, Lu YC, Kapse K, Jacobs M, Andescavage N, Donofrio MT, et al. In Utero MRI Identifies Impaired Second Trimester Subplate Growth in Fetuses with Congenital Heart Disease. Cereb Cortex. 2021 Oct 28;bhab386.

23. Andescavage N, Yarish A, Donofrio M, Bulas D, Evangelou I, Vezina G, et al. 3-D volumetric MRI evaluation of the placenta in fetuses with complex congenital heart disease. Placenta. 2015 Sept;36(9):1024–30.

24. Matthiesen NB, Henriksen TB, Agergaard P, Gaynor JW, Bach CC, Hjortdal VE, et al. Congenital Heart Defects and Indices of Placental and Fetal Growth in a Nationwide Study of 924□422 Liveborn Infants. Circulation. 2016 Nov 15;134(20):1546–56.

25. Rychik J, Goff D, McKay E, Mott A, Tian Z, Licht DJ, et al. Characterization of the Placenta in the Newborn with Congenital Heart Disease: Distinctions Based on Type of Cardiac Malformation. Pediatr Cardiol. 2018 Aug;39(6):1165–71.

26. Snoep MC, Aliasi M, van der Meeren LE, Jongbloed MRM, DeRuiter MC, Haak MC. Placenta morphology and biomarkers in pregnancies with congenital heart disease – A systematic review. Placenta. 2021 Sept;112:189–96.

27. Cromb D, Slator PJ, Hall M, Price A, Alexander DC, Counsell SJ, et al. Advanced magnetic resonance imaging detects altered placental development in pregnancies affected by congenital heart disease. Sci Rep. 2024 May 29;14(1):12357.

28. Albalawi A, Brancusi F, Askin F, Ehsanipoor R, Wang J, Burd I, et al. Placental Characteristics of Fetuses With Congenital Heart Disease. J Ultrasound Med. 2017;36(5):965–72.

29. Miremberg H, Gindes L, Schreiber L, Raucher Sternfeld A, Bar J, Kovo M. The association between severe fetal congenital heart defects and placental vascular malperfusion lesions. Prenat Diagn. 2019 Oct;39(11):962–7.

30. Leon RL, Sharma K, Mir IN, Herrera CL, Brown SL, Spong CY, et al. Placental vascular malperfusion lesions in fetal congenital heart disease. Am J Obstet Gynecol. 2022 May 21;S0002-9378(22)00389-1.

31. Schlatterer SD, Murnick J, Jacobs M, White L, Donofrio MT, Limperopoulos C. Placental Pathology and Neuroimaging Correlates in Neonates with Congenital Heart Disease. Sci Rep. 2019 Mar 11;9:4137.

32. Nijman M, van der Meeren LE, Nikkels PGJ, Stegeman R, Breur JMPJ, Jansen NJG, et al. Placental Pathology Contributes to Impaired Volumetric Brain Development in Neonates With Congenital Heart Disease. J Am Heart Assoc. 2024 Mar 5;13(5):e033189.

33. Thilaganathan B. Preeclampsia and Fetal Congenital Heart Defects: Spurious Association or Maternal Confounding? Circulation. 2017 July 4;136(1):49–51.

34. Cohen JA, Rychik J, Savla JJ. The placenta as the window to congenital heart disease. Curr Opin Cardiol. 2021 Jan;36(1):56–60.

35. Leon RL, Mir IN, Herrera CL, Sharma K, Spong CY, Twickler DM, et al. Neuroplacentology in congenital heart disease: placental connections to neurodevelopmental outcomes. Pediatr Res. 2021 Apr 16;1–8.

36. Snoep MC, Bet BB, Zwanenburg F, Knobbe I, Linskens IH, Pajkrt E, et al. Factors Related to Fetal Demise in cases with Congenital Heart Defects. Am J Obstet Gynecol MFM. 2023 Aug;5(8):101023.

37. Cromb D, Slator PJ, De La Fuente M, Price AN, Rutherford M, Egloff A, et al. Assessing within-subject rates of change of placental MRI diffusion metrics in normal pregnancy. Magn Reson Med. 2023 May 15;90(3):1137–50.

38. Aviles Verdera J, Payette K, Hall M, Bradshaw C, Story L, Bansal S, et al. Comprehensive assessment of uterine contractility using a large database of dynamic T2∗ studies. Placenta. 2025 Sept 1;169:39–48.

39. Cox JL, Holden JM, Sagovsky R. Detection of Postnatal Depression: Development of the 10-item Edinburgh Postnatal Depression Scale. Br J Psychiatry. 1987 June;150(6):782–6.

40. Spielberger CD, Gorsuch RL. Manual for the state-trait anxiety inventory (form Y)□:. Consulting Psychologists Press; 1983.

41. Block AJ, McQuillen PS, Chau V, Glass H, Poskitt KJ, Barkovich AJ, et al. Clinically silent preoperative brain injuries do not worsen with surgery in neonates with congenital heart disease. J Thorac Cardiovasc Surg. 2010 Sept;140(3):550–7.

42. Bergink V, Kooistra L, Lambregtse-van den Berg MP, Wijnen H, Bunevicius R, van Baar A, et al. Validation of the Edinburgh Depression Scale during pregnancy. J Psychosom Res. 2011 Apr;70(4):385–9.

43. Levis B, Negeri Z, Sun Y, Benedetti A, Thombs BD, DEPRESsion Screening Data (DEPRESSD) EPDS Group. Accuracy of the Edinburgh Postnatal Depression Scale (EPDS) for screening to detect major depression among pregnant and postpartum women: systematic review and meta-analysis of individual participant data. BMJ. 2020 Nov 11;371:m4022.

44. Grant KA, McMahon C, Austin MP. Maternal anxiety during the transition to parenthood: A prospective study. J Affect Disord. 2008 May 1;108(1):101–11.

45. Yan J, Fine J. Estimating equations for association structures. Stat Med. 2004 Mar 30;23(6):859–74; discussion 875-877,879-880.

46. Højsgaard S, Halekoh U, Yan J. The R Package geepack for Generalized Estimating Equations. J Stat Softw. 2006;15:1–11.

47. Xu Z, Fine JP, Song W, Yan J. On GEE for Mean-Variance-Correlation Models: Variance Estimation and Model Selection. Stat Med. 2025 Jan 15;44(1–2):e10271.

48. Melbourne A, Aughwane R, Sokolska M, Owen D, Kendall G, Flouri D, et al. Separating fetal and maternal placenta circulations using multiparametric MRI. Magn Reson Med. 2019 Jan;81(1):350–61.

49. Melbourne A. On the use of multicompartment models of diffusion and relaxation for placental imaging. Placenta. 2021 Sept 1;112:197–203.

50. Seth S, Lewis AJ, Saffery R, Lappas M, Galbally M. Maternal Prenatal Mental Health and Placental 11β-HSD2 Gene Expression: Initial Findings from the Mercy Pregnancy and Emotional Wellbeing Study. Int J Mol Sci. 2015 Nov 17;16(11):27482–96.

51. Galbally M, Watson SJ, Lappas M, de Kloet ER, van Rossum E, Wyrwoll C, et al. Fetal programming pathway from maternal mental health to infant cortisol functioning: The role of placental 11β-HSD2 mRNA expression. Psychoneuroendocrinology. 2021 May;127:105197.

52. Meyer JS, Novak MA. Assessment of prenatal stress-related cortisol exposure: focus on cortisol accumulation in hair and nails. Dev Psychobiol. 2021 Apr;63(3):409–36.

53. Hutter J, Ho A, Jackson LH, Slator PJ, Chappell LC, Hajnal JV, et al. An efficient and combined placental T1 -ADC acquisition in pregnancies with and without pre-eclampsia. Magn Reson Med. 2021 Nov;86(5):2684–91.

54. Lu T, Wang Y, Guo A, Wei C, Chen Y, Wang S, et al. Standard diffusion-weighted, diffusion kurtosis and intravoxel incoherent motion MR imaging of the whole placenta: a pilot study of volumetric analysis. Ann Transl Med. 2022 Mar;10(6):269.

55. Lahti-Pulkkinen M, Cudmore MJ, Haeussner E, Schmitz C, Pesonen AK, Hämäläinen E, et al. Placental Morphology Is Associated with Maternal Depressive Symptoms during Pregnancy and Toddler Psychiatric Problems. Sci Rep. 2018 Jan 15;8(1):791.

56. Giménez Y, González E, Fatjó F, Mallorquí A, Hernández S, Arranz A, et al. Anxiety and depression during pregnancy: Differential impact in cases complicated by preeclampsia and preterm premature rupture of membranes. PLOS ONE. 2025 Apr 29;20(4):e0302114.

57. Edwards AD, Redshaw ME, Kennea N, Rivero-Arias O, Gonzales-Cinca N, Nongena P, et al. Effect of MRI on preterm infants and their families: a randomised trial with nested diagnostic and economic evaluation. Arch Dis Child Fetal Neonatal Ed. 2018 Jan;103(1):F15–21.

58. Wu Y, Kapse K, Jacobs M, Niforatos-Andescavage N, Donofrio MT, Krishnan A, et al. Association of Maternal Psychological Distress With In Utero Brain Development in Fetuses With Congenital Heart Disease. JAMA Pediatr. 2020 Mar 1;174(3):e195316.

59. Optimizing Psychological Health Across the Perinatal Period: An Update on Maternal Cardiovascular Health: A Scientific Statement From the American Heart Association | Journal of the American Heart Association. 2025 Mar 4;14(5):e041369.

