## Supplementary Materials for "Maternal prenatal stress is associated with altered placental microstructure in low-risk pregnancies and pregnancies with Congenital Heart Disease"

### **Supplementary Material**

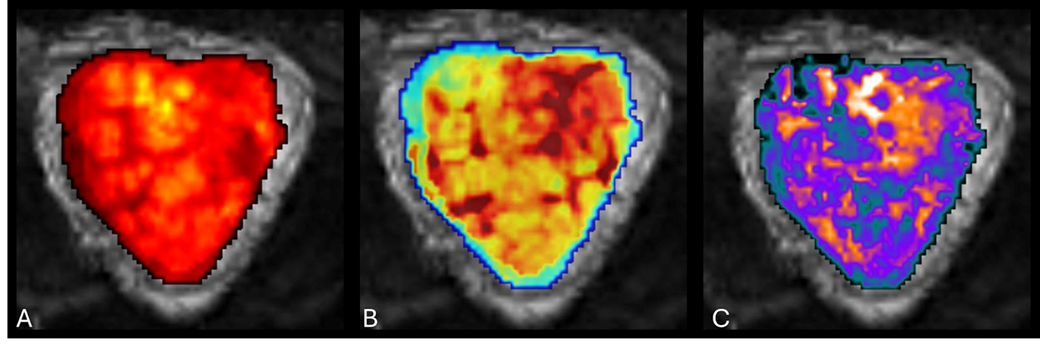

**Supplementary Figure 1.** A mid-parenchymal single-slice from placental T2* (A) and ADC (B) maps overlaid on acquired placental diffusion-relaxation MRI data for a healthy control participant scanned at 27^+5^ weeks.

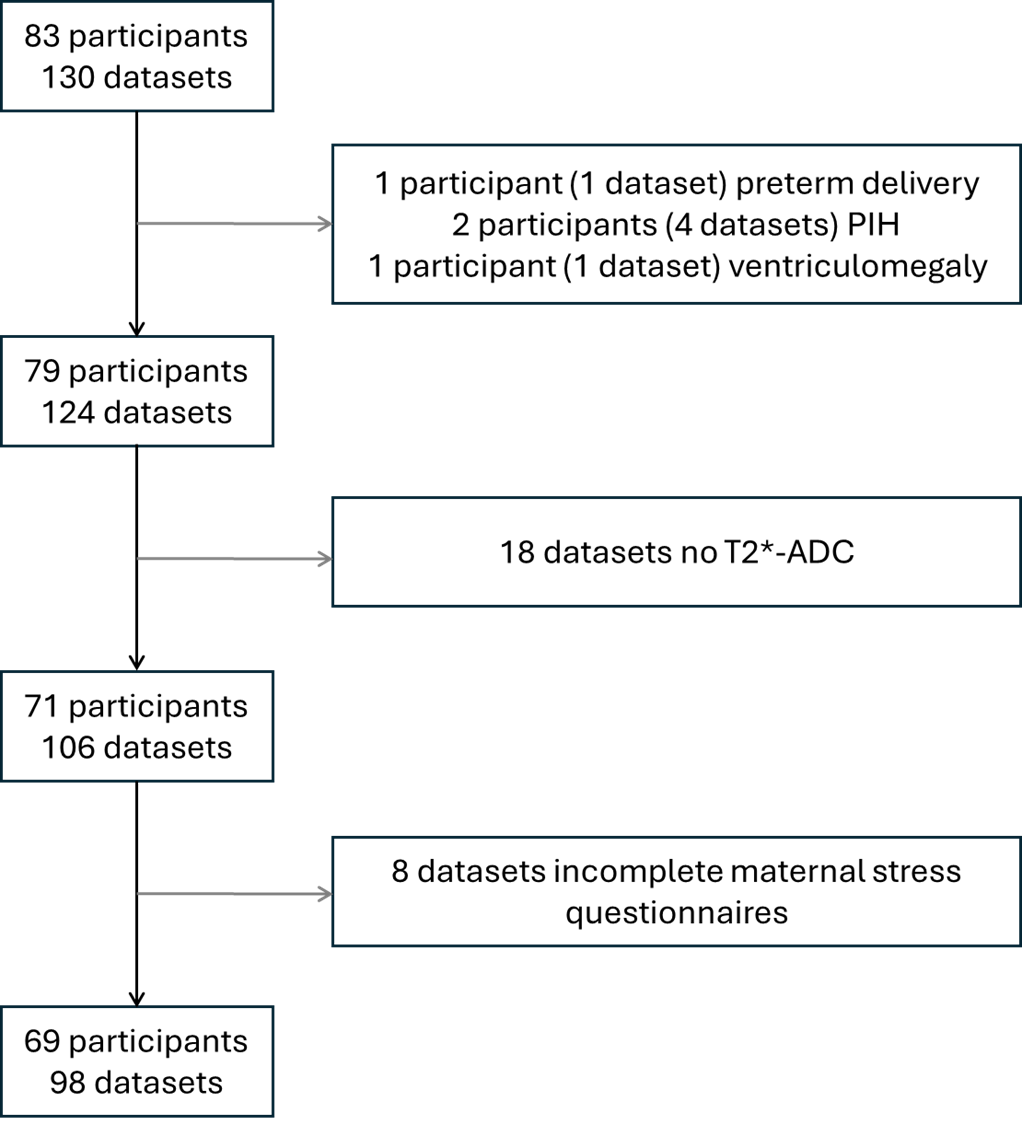

**Supplementary Figure 2.** Diagram of participant exclusions. PIH= pregnancy-induced hypertension

**Supplementary Table 1.** Combined T2*-ADC multi-echo gradient-echo placental MRI scan acquisition parameters

Orientation: Coronal plane to maternal habitus, FOV = 300 × 320 × 84 mm

Resolution: 3 mm^3^ isotropic

Echo Time = (78, 114, 150, 186) ms, Repetition Time = 7.5 ms, SENSE factor = 2.5

b = (5, 10, 25, 50, 100, 200, 400, 600, 1200, 1600) s mm^−2^; 3 directions

b = 18 s mm^−2^; 8 directions

b = 36 s mm^−2^; 7 directions

b = 800 s mm^−2^; 15 directions

| **Supplementary Table 2.** Moderating effect of CHD diagnosis on relationship between maternal stress measures and placental microstructure and function. | | | |
| --- | --- | --- | --- |
| Maternal Stress Measure | β (SE) | Wald statistic | P-value (p_FWE_) |
| Placental ADC | | | |
| EPDS maternal depression*CHD | -0.121 (0.155) | 0.600 | 0.438 (0.576) |
| STAI state anxiety*CHD | -0.354 (0.151) | 5.54 | 0.019 (0.114) |
| STAI trait anxiety*CHD | -0.231 (158) | 2.13 | 0.144 (0.576) |
| Placental T2* | | | |
| EPDS maternal depression*CHD | -0.204 (0.142) | 2.06 | 0.151 (0.576) |
| STAI state anxiety*CHD | -0.286 (0.138) | 3.30 | 0.038 (0.190) |
| STAI trait anxiety*CHD | -0.163 (0.128) | 1.61 | 0.204 (0.576) |
| Analyses adjusted for other maternal stress measures, previous mental health treatment, male sex, GA at scan, GA at scan^2^. ADC= Apparent diffusion coefficient, EPDS= Edinburgh Postnatal Depression Scale, STAI= State-Trait Anxiety Inventory | | | |
